# Genetic ancestral patterns in *CYP2D6* alleles: structural variants, rare variants, and clinical associations in 479,144 UK Biobank genomes

**DOI:** 10.1101/2024.07.23.24310892

**Authors:** Xiao Jiang, Fengyuan Hu, Xueqing Zoe Zou, Ali Abbasi, Sri V. V. Deevi, Santosh S. Atanur, Amanda O’Neill, Jen Harrow, Margarete Fabre, Quanli Wang, Slavé Petrovski, William Rae, Oliver Burren, Katherine R. Smith

## Abstract

Cytochrome P450 2D6 (*CYP2D6*) is involved in metabolising over 20% of clinical drugs, yet its genetic variation across ancestries is underexplored in large-scale whole-genome sequencing (WGS) datasets. We analysed WGS data from 479,144 UK Biobank participants, identifying 95 distinct *CYP2D6* star alleles across five biogeographic groups. Of these, 48 alleles had currently unknown effects. These alleles were more prevalent in African, admixed American, and South Asian groups (∼5%) compared to European and East Asian groups (∼2%), affecting the ability to provide pharmacogenomics recommendations across ancestries. We identified 99,656 (20.8%) individuals carrying *CYP2D6* structural variations and predicted the *CYP2D6* ultra-rapid metaboliser phenotype to be most common in Africans (4.5%) and rarest in East Asians (0.32%). Less than half (45.7%) of rare protein-truncating variant carriers were categorised as poor or intermediate metabolisers, indicating an underrepresentation of rare functional variants in the current *CYP2D6* star allele evaluation. Phenome-wide association studies confirmed links with narcotic allergies and found new associations with plasma BAFFR and BAFF proteins, offering insights for the BAFF-targeted clinical therapy. Collectively, this largest WGS study of *CYP2D6* to date highlights the importance of leveraging all genetic variations for pharmacogenomic insights affecting therapeutic safety and development.

## Introduction

Pharmacogenomics (PGx) explores the influence of the genome on an individual’s response to medication. Consequently, PGx plays a pivotal role in advancing the principles and practices of precision medicine by utilising genetic characteristics to optimise drug therapy and enhance treatment outcomes. Pharmacogenes, are genes heavily involved in drug metabolism. For example, cytochrome P450 2D6 (*CYP2D6*) is responsible for metabolising over 20% of currently clinically prescribed drugs, particularly those with psychoactive properties^1^. *CYP2D6* is highly polymorphic, with over 160 haplotype patterns documented in the Pharmacogenomics Knowledgebase (PharmGKB)^2^. Referred to as star alleles, these encompass combinations of small nucleotide variants (SNVs), small in-frame insertions and deletions (INDELs), and structural variants (SVs)^3^, including hybrid gene arrangements with its upstream adjacent paralogous pseudogene, *CYP2D7*^4,5^. Additionally, despite of limited cohort sizes for non-European genetic ancestries, it has been shown that broad genetic ancestral groups exhibit distinctive *CYP2D6* star allele frequencies^6–10^.

Pharmacogenomic resources predict an individual’s *CYP2D6* enzyme activity level, or gPheno based on their star allele haplotypes, with five potential metaboliser classifications: poor (PM), intermediate (IM), normal (NM), ultra-rapid (UM) or unknown^11^. The *CYP2D6* enzyme activity can be associated with either poor efficacy or adverse events when specific pharmacogene-metabolised therapeutic treatments are administered^12^. Therefore, gPheno prediction can be critical in clinical decision-making for prescribing drugs known to be metabolised by *CYP2D6*. The Clinical Pharmacology Implementation Consortium (CPIC)^13^ has published clinical guidelines for several *CYP2D6*-metabolised drugs, including Atomoxetine^14^, Ondansetron and Tropisetron^15^, Tamoxifen^16^, Tricyclic Antidepressants^17^, and certain types of Opioids^18^. Recently, studies have evaluated *CYP2D6* genotypes for the UK Biobank (UKBB) participants, integrating imputed genotypes with whole-exome-sequencing data^19,20^. However, the lack of SV calls in these studies meant that certain loss of function star alleles such as *5, *13, and *36 could not be detected, impacting allele frequency estimation and therefore gPheno prediction.

In this study, we used the DRAGEN (v3.7.8) *CYP2D6* genotype caller^21^ to identify the *CYP2D6* star allele genotypes for 490,524 multi-ancestry UKBB participants using whole-genome-sequencing (WGS) data^22^. We used these data to characterise the genetic architecture of *CYP2D6* across five broad genetic ancestries and assess the associations of *CYP2D6* gPheno predictions with 20,488 clinical and molecular phenotypes. This study, the largest WGS analysis of *CYP2D6* to date, highlights the importance of leveraging all types of genetic variation in a large population-based cohort to provide PGx insights that may impact therapeutic safety and development.

## Methods

### UKBB whole genome Sequencing processing

Whole-genome-sequencing (WGS) data of the UKBB participants were generated by deCODE Genetics and the Wellcome Trust Sanger Institute as part of a public-private partnership involving AstraZeneca, Amgen, GlaxoSmithKline, Johnson & Johnson, Wellcome Trust Sanger, UK Research and Innovation, and the UKBB. The WGS sequencing methods and QC have been previously described^22,23^. Briefly, genomic DNA underwent paired-end sequencing on Illumina NovaSeq6000 instruments with a read length of 2×151 and an average coverage of 32.5x. Conversion of sequencing data in BCL format to FASTQ format and the assignments of paired-end sequence reads to samples were based on 10-base barcodes, using bcl2fastq v2.19.0. Initial quality control was performed by deCODE and Wellcome Sanger, which included sex discordance, contamination, unresolved duplicate sequences, and discordance with microarray genotyping data checks.

### AstraZeneca Centre for Genomics Research (CGR) UKB WGS small variant, *CYP2D6* star allele, and copy number variant calling

UKBB genomes were processed at AstraZeneca CGR using the provided CRAM format files. A custom-built Amazon Web Services (AWS) cloud compute platform running Illumina DRAGEN Bio-IT Platform Germline Pipeline v3.7.8 (DRAGEN v3.7.8) was used to align the reads to the GRCh38 genome reference and to call small variants. Small variants were annotated using SnpEff v4.3^24^ against Ensembl Build 38.92^25^. We adopted the DRAGEN v3.7.8 in-build *CYP2D6* Caller employing the method in Cyrius^21^ to identify the *CYP2D6* star allele diplotype for each UKBB genome. We retained for downstream analysis calls from 482,033 individuals where the caller could determine the two haplotypes with high confidence (status “PASS”), discarding those where the variants could not be matched to star alleles (“No_call”, n=3,594) or were consistent with more than one possible genotype (“More_than_one_possible_genotype”, n=4,931). Furthermore, we used *Peddy*^26^ and 1000 Genomes Project data^27,28^ to classify participants (peddy probability ≥ 0.80) into broad genetic ancestries. Copy number variants of UKBB WGS were called by DRAGEN v3.7.8 germline CNV caller. Post-hoc sample QC and variant QC were applied to ensure a good quality CNV call set (median mendelian violation rate = 4.1%; heterozygous de novo rate = 1.9%). According to the overlapping between CNVs and genes, we annotated the functional consequences of CNVs against 19,348 protein-coding genes from Ensembl Build 38.92^25^. All genes, exons, UTRs from Ensembl Build 38.92 annotation were included in the analyses. If a gene has multiple transcripts, we included all transcripts in the analyses. UTRs were defined as the union regions of all transcripts. Promoters were defined as the 1kb window before the transcription start site.

### Mendelian Consistency in the UKB trios and monozygotic twins

We utilised KING v2.2.7 on the UKBB array genotypes with parameters “--build --degree 1” and determined 1,047 sets of trios (consisting of two parents and an offspring) within the UKBB WGS dataset. Among these, 1,014 trios had *CYP2D6* star allele genotypes called for all three members. Mendelian consistency within trios was determined by confirming that the offspring inherited one haplotype star allele from each parent. Additionally, we identified 362 pairs of monozygotic twins from the UKBB WGS data, with *CYP2D6* star allele genotypes called for both individuals in 360 of these pairs. Mendelian consistency in monozygotic twins was defined by the presence of identical *CYP2D6* star allele genotypes in both individuals.

### *CYP2D6* star allele haplotype frequency estimation

The star allele frequency was assessed separately within each broad genetic ancestral group in the UKBB dataset. Within each group, we initially tallied the presence of each haplotype. Instances of identical gene duplications with different copies were evaluated individually, except for those with three or more copies of *1, *2, *4, where counts were aggregated and categorised as “*1x≥3”, “*2x≥3”, and “*4x≥3” to facilitate comparison with data in PharmGKB^29^. Subsequently, the frequency for each star allele haplotype was estimated by dividing the haplotype count by the total number of haplotypes within the respective ancestral groups (i.e., twice the sample size). The estimated frequencies of star allele haplotypes in the AFR, AMR, EAS, EUR, and SAS groups were compared with those of “African American/Afro-Caribbean”, “American”, “East Asian”, “European”, and “Central/South Asian” in PharmGKB using the *cor.test()* function within the “stats” package (v4.3.3) in R.

### *CYP2D6* genetically predicted metaboliser phenotypes

The activity values for the identified star alleles were sourced from the functionality table of *CYP2D6* alleles in PharmGKB^30^. To estimate the *CYP2D6* metaboliser activity score for each individual, we summed the activity values corresponding to both their star allele haplotypes. For star alleles with predicted duplications or higher-order multiplications, we multiplied the activity value by the number of repetitions. Following the definition provided by CPIC^11,31^, individuals were categorised as follows: 1) *CYP2D6* poor metaboliser (PM) if their activity score equalled zero; 2) *CYP2D6* intermediate metaboliser (IM) if their activity score fell between 0 and 1.25 (exclusive); 3) *CYP2D6* normal metaboliser (NM) if their activity score ranged from 1.25 to 2.25 (inclusive); 4) *CYP2D6* ultra-rapid metaboliser (UM) if their activity score exceeded 2.25 (exclusive); 5) *CYP2D6* indeterminate if they carried a star allele of unknown function. In addition, 23 individuals who carrying unknown effect star alleles but with an activity score greater than 2.25 based on remaining alleles were classified as UM metabolisers.

### *CYP2D6* predicted protein-truncating variation definition and filtration criteria

As described in the previous study^32^, we defined protein-truncating variants (PTVs) based on SnpEff v4.3^24^ annotations of variants as exon_loss_variant, frameshift_variant, start_lost, stop_gained, stop_lost, splice_acceptor_variant, splice_donor_variant, gene_fusion, bidirectional_gene_fusion, rare_amino_acid_variant, and transcript_ablation.

The applied quality control filters were as follows: a minor allele frequency (MAF) of ≤ 5%, a minimum coverage of 10X, and annotation within CCDS transcripts (release 22; approximately 34 Mb). For homozygous genotypes, alternate reads were limited to 80%, while for heterozygous variants, the proportion of alternate reads had to be between 0.25 and 0.8. Additionally, a binomial test for the alternate allele proportion in the heterozygous state had to show no significant departure from 50% (P > 1 × 10^-6^). Other criteria included a genotype quality (GQ) of ≥ 20, a Fisher’s strand bias score (FS) of ≤ 200 for indels and ≤ 60 for SNVs, a mapping quality (MQ) of ≥ 40, a quality score (QUAL) of ≥ 30, a read position rank sum score (RPRS) of ≥ -2, and a mapping quality rank sum (MQRS) of ≥ -8. Variants had to pass the DRAGEN variant status. The variant site must have at least 10X coverage in over 90% of sequences and must not fail any of the specified quality control criteria in 5% or more of the sequences. Furthermore, the variant site required tenfold coverage in at least 25% of gnomAD exomes^33^, and if present in gnomAD exomes, the variant needed an exome z-score of ≥ -2.0 and an exome MQ of ≥ 30.

### Statistical Associations

We conducted ancestry-specific association tests for 12 predefined contrasts of *CYP2D6* metaboliser categories for 15,909 binary clinical outcomes and 1,656 quantitative traits collected from the UKBB dataset. We tested 11 contrasts defined so that the predicted activity score in cases was higher than controls, and one negative control contrast comparing extreme (PM and UM) to central (IM and NM) categories. We adopted Fisher’s exact test for the association analyses against the binary clinical outcomes, and linear regression test for quantitative traits, which were first transformed by rank-based inverse normalisation. The linear regression was adjusted for the age at recruitment and the genetically predicted sex. Subsequently, a meta-analysis was conducted to consolidate the results of the tested phenotypes within each ancestral group. For combining the association results of binary clinical outcomes, we employed the exact conditional test using the *mantelhaen.test()* within the “stats” package (v4.3.3) in R, while the association results of quantitative traits were combined using the fixed-effect inverse variance weighting method. Due to the limited numbers of non-EUR participants with UKBB plasma protein abundance data currently available, we performed a pan-ancestry analysis by regressing the *CYP2D6* metaboliser status for 47,599 individuals against 2,941 plasma protein abundance levels corresponding to 2,923 proteins, adopting the same adjustments described previously^34^. To address multiple testing, we implemented the Bonferroni correction accounting for the number of tested phenotypes and models, setting the corrected significance level at 2.0×10^-7^.

### Test for Heterogeneity

In comparison to individuals classified as *CYP2D6* NMs, those identified as PMs and IMs exhibited significant associations with plasma protein abundance of POMC, BAFF and BAFFR. We assessed evidence for heterogeneity in associations with these proteins for PMs vs NMs and IMs vs NMs, or between PMs vs IMs and PMs vs NMs. For example, to test for heterogeneity between PMs vs NMs and IMs vs NMs, we first ensured the independence of associations between the two pairs of *CYP2D6* metaboliser comparisons by dividing *CYP2D6* NM individuals randomly into two subgroups based on the ratio of PM and IM cohort sizes (NM_P_ and NM_I_). Subsequently, we conducted regression analyses for the newly formed metaboliser pairs (PM vs. NM_P_; IM vs. NM_I_) against POMC, BAFF and BAFFR, respectively, employing the same adjustments as previously reported^34^. We then evaluated the heterogeneity of associations using the *rma()* function (method=“REML”) within the “metafor” package (v4.4.0) in R. This process was repeated 1,000 times, and the median of the P-values for Cochran’s Q test was reported. We defined significance of heterogeneity as Cochran’s Q test P-value < 5%.

## Results

### *CYP2D6* genotyping and phenotyping calling

Excluding 36 individuals under consent withdrawals, for the remaining 490,524 UKBB genomes, we were able to call *CYP2D6* star allele genotypes for 481,999 (98.3%) individuals, with additional 1% ambiguous genotype calls (“More than one possible genotype”, N=4,931) and 0.7% “No Call” status (N=3,594), returned by DRAGEN v3.7.8. For successful calls, we examined 1,014 trios and 360 monozygotic twins and found Mendelian consistency rates (**Methods**) of 98.8% and 100% respectively, comparable with that reported previously from 1000 Genome (1KG) Project^21^. Notably, the 12 families that demonstrated Mendelian inconsistency encompassed at least one *CYP2D6* SV carrier. We used 1KG data to classify individuals in to five broad genetic ancestral groups: African (AFR; N=8,822), admixed American (AMR; N=955), East Asian (EAS; N=2,470), European (EUR; N=456,778), and South Asian (SAS; N=10,119) (**Methods**). Subsequently, we used star allele genotypes to predict an individual’s *CYP2D6* activity and overall *CYP2D6* metaboliser phenotype (gPheno). In total, 479,144 participants with *CYP2D6* gPhenos were available for downstream analyses (**Figure 1**).

**Figure 1.**
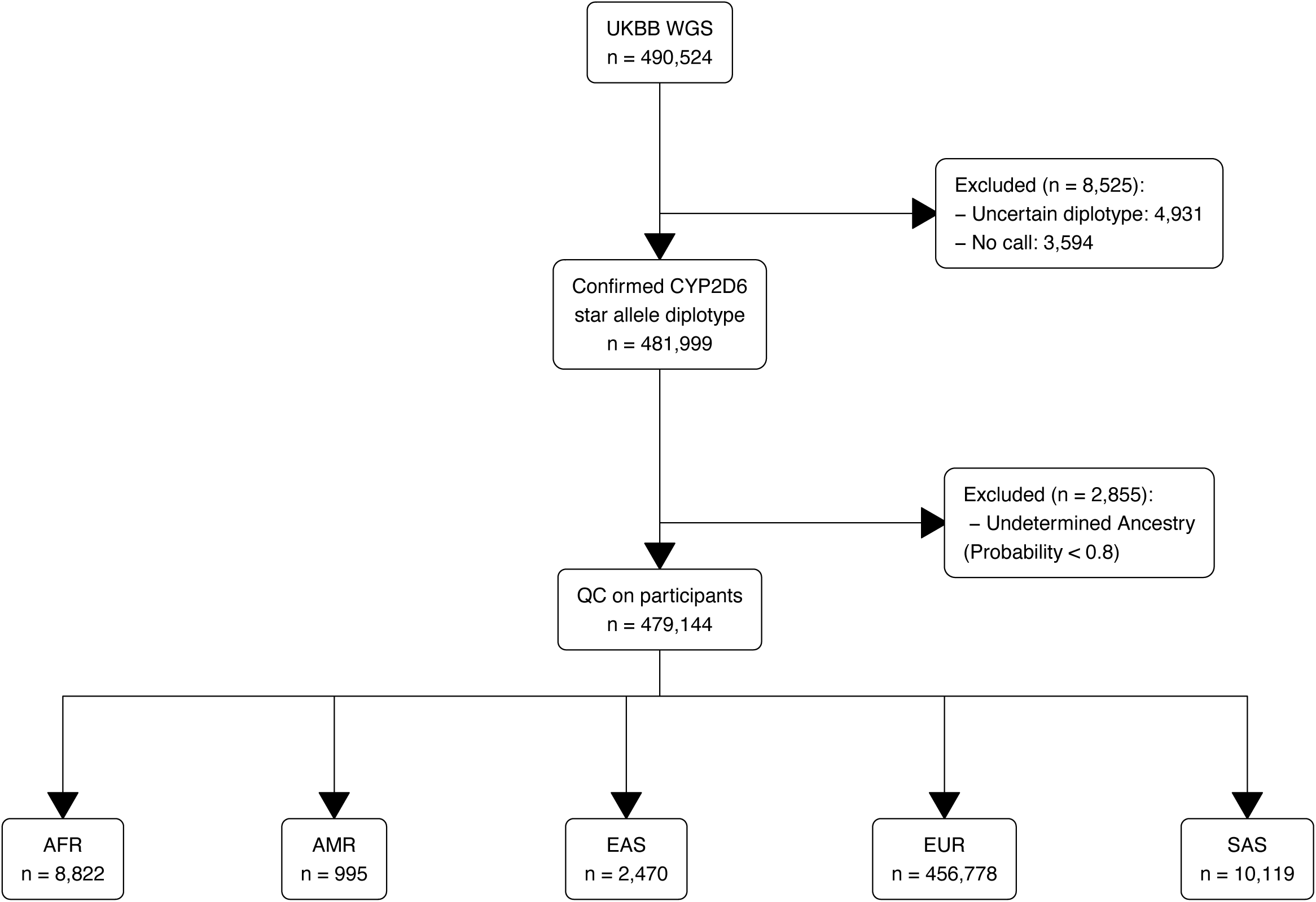
Overview of *CYP2D6* star allele calling workflow and sample filtrations in the UKBB.

Utilising a standalone copy number variant (CNV) calling pipeline (**Methods**), we identified nine *CYP2D6* CNVs spanning the *CYP2D8P-CYP2D7*-*CYP2D6* genomic region (GRCh38 chr22:42,126,499-42,155,001), encompassing 10,373 participants in the UKBB. Among these, 9,956 individuals were involved into our study, with 9,940 (99.8%) being classified as SV carriers based on the identified *CYP2D6* star alleles as well (**Supplementary Table 1**).

### Underrepresentation of diversity in the current evaluation of *CYP2D6* genotypes

In total, we identified 95 distinct *CYP2D6* star alleles. These star alleles were unevenly distributed among the five broad genetic ancestry groups (**Figure 2A**, **Supplementary Table 2**), with the proportions of both non-functional (activity value=0) and normal functioning (activity value=1) star alleles being the smallest in the EUR individuals. Notably, of the 95 star alleles, 48 (50.5%) lacked known effects on *CYP2D6* enzyme activities according to PharmGKB. Overall, we found these constituted between 30% to 50% of star alleles identified across the genetic ancestries examined (**Figure 2A**). Although indeterminate star alleles were only observed in 1.9% of UKBB participants, they led to challenges in gPheno classification for, on average 5% of AFR, AMR and SAS individuals in contrast to 2% of EUR and EAS.

**Figure 2.**
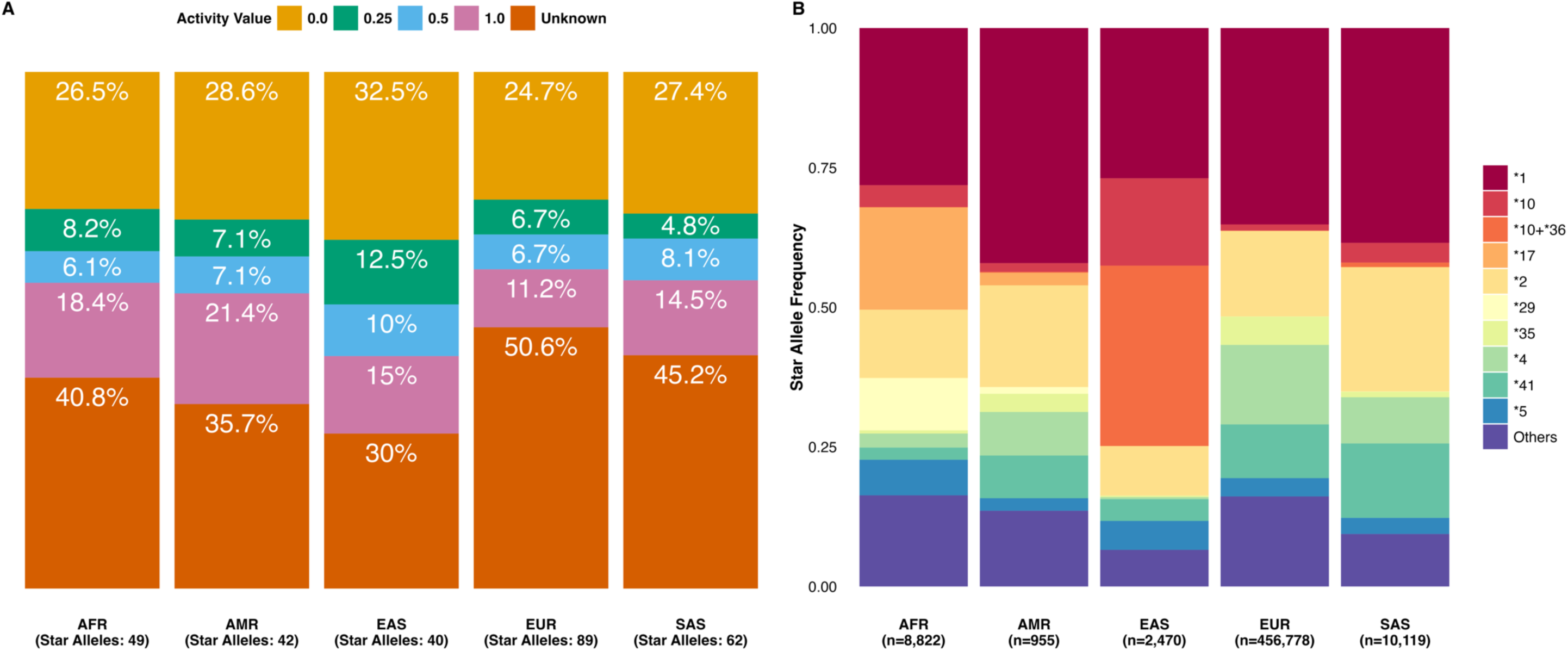
Overview of distributions of *CYP2D6* star allele called in the UKBB. A) Distribution of functionality values of the identified *CYP2D6* star alleles in each genetically predicted ancestral group. The x-axis includes the count of distinct star alleles identified within each ancestral group. Each bar was segmented and coloured according to the distribution of star alleles based on their respective functionality values. **B) Distribution of the most prevalent (top 5) star alleles of each ancestry and their frequencies in other ancestries.** The x-axis provides the sample size for each ancestral group. Ten star alleles are highlighted, of which, two were SVs (*5, *10+*36). There were three normal-function alleles (activity value=1; *1, *2, *35), five decreased-function alleles (activity value= 0.25 or 0.5; *10, *10+*36, *17, *29, *41), and two non-functional alleles (activity value=0; *4, *5).

In total, we were able to predict gPhenos for 424,682 individuals (88.6%) whose star allele diplotype, activity scores, and the corresponding predicted phenotype were documented in PharmGKB, where all predicted metaboliser phenotypes aligned consistently with the *CYP2D6* gPhenos within our study. The remaining 54,462 individuals (11.4%) all carried non-identical SVs with duplications or higher-order multiplications (e.g., *10+*36, *4+*68). We further conducted genetic ancestry-specific analysis to assess the consistency of *CYP2D6* star allele haplotype frequencies to those reported in PharmGKB (**Methods; Supplementary** Figure 1**; Supplementary Table 3**). Overall, we observed a high degree of correlation in haplotype frequencies among four genetic ancestries: AFR (r=0.97), AMR (r=0.99), EUR (r=0.98), and SAS (r=0.97). Notably, the correlation in individuals with EAS was lower (r=0.86). This was primarily due to the absence of frequency information for the *10+*36 star allele combination in PharmGKB as we were able to improve correlation to 0.89 by incorporating information from a Chinese- and Malay-focused cohort^10^. As previously reported^10^, we observed a lower frequency of *10 within EAS individuals with respect that reported by the PharmGKB. We also compared star allele frequencies of the UKBB genetic African ancestry with those of “African American/Afro-Caribbean” and “Sub-Saharan African” documented in the PharmGKB (**Supplementary** Figure 2**; Supplementary Table 4**).

We selected the five most common *CYP2D6* star allele haplotypes within each genetic ancestry and then compared their prevalence across the remaining genetic ancestry groups (**Figure 2B**). This highlighted ten distinct star alleles, including two structural variants (*CYP2D6*\*5, *10+*36). Among these ten star alleles, three were normal-functioning alleles (activity value=1) of which two were common across ancestries (*1, *2), with the other (*35), found to be more common in EUR (5.1%) and AMR (3.2%) individuals. Five were reduced-functional alleles (activity value=0.25 or 0.5) including *17 and *29 that were over nine-times more frequent in AFR individuals than other genetic ancestries. The *10 and*10+*36 hybrid arrangements were most frequent in EAS individuals (15.6% and 32.3%, respectively); whereas the remaining *41 had a significant enrichment in the SAS individuals. Finally, the two remaining star alleles were non-functional (activity value=0). These included the EUR-enriched *4 (14.2%) and the whole gene deletion variation *5 most frequently found in AFR and EAS individuals (**Figure 2B; Supplementary Table 3**).

### Evaluation of *CYP2D6* structural variations

*CYP2D6* SVs are essential for precisely determining an individual’s *CYP2D6* gPheno, particularly for the UM category. These *CYP2D6* SVs can include whole gene deletions (i.e., *5), duplications or higher-order multiplications of identical gene copies (e.g., *1×2, *4×2) and non-identical gene copies (e.g., *4+*68, *10+*36), singleton hybrid genes (e.g., *4.013, *13), and combined arrangements (e.g., *4+*68×2, *10+*36×2). We found that 99,656 (20.8%) of participants carried at least one *CYP2D6* SV, and that these individuals were spread unevenly across the five broad genetic ancestry groups (**Figure 3A**). Notably, 63.6% EAS individuals carried one or more *CYP2D6* SVs, a figure higher than a previous estimate of 55.6% in an admixed Asian population from Singapore^10^. To investigate this, we randomly selected a comparable cohort size from UKBB individuals (N=2,000) with proportions of EAS and SAS individuals matched to those in the Singaporean study. By 1,000-time iterations, we observed a 58.3% [IQR 57.9-58.7%] median SV carrying frequency. Therefore, we conclude that the observed discrepancy (63.6% vs 55.6%) is most likely due to differences in admixture between the two studies.

**Figure 3.**
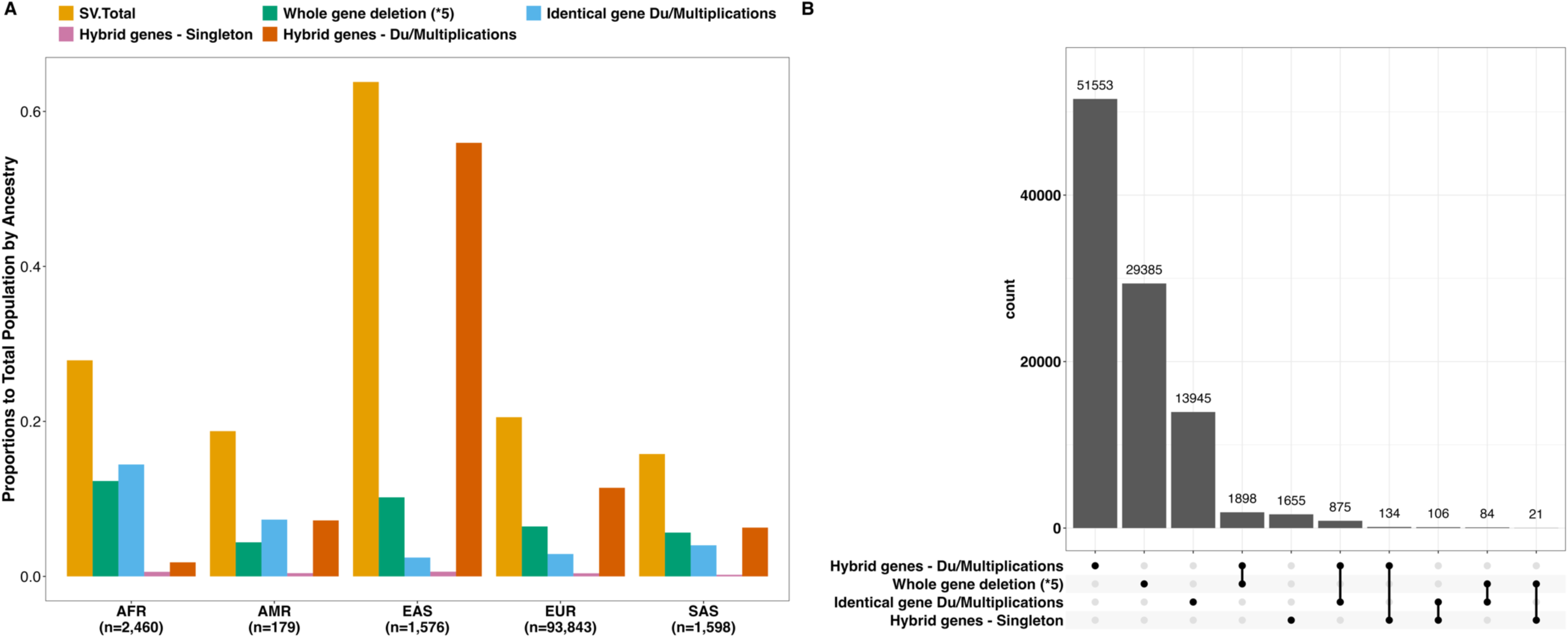
Overview of distributions of *CYP2D6* SVs in the UKBB. A) Distributions of *CYP2D6* SVs and the subtypes across genetic ancestries. The x-axis displays the count of *CYP2D6* SV carriers within each ancestral group. The proportions on the y-axis were determined by dividing the carrier count for each SV type by the total sample size of the respective ancestral population in the UKBB. B) Distribution of carriers of each *CYP2D6* SV type and intersection of subtypes. Over half of *CYP2D6* SV carriers (54.6%) were carrying the non-identical gene duplications or higher-order multiplications. Notably, there were 2,003 individuals carrying both the whole gene deletion (*5) and another *CYP2D6* SV type.

#### Gene deletions

*CYP2D6*\*5 denotes a complete deletion of the *CYP2D6* gene, resulting in a total loss of enzyme activity. Overall, it was one of the most common SVs with 31,388 (31.5% of the total *CYP2D6* SV carriers) carriers and consistent with a previous report^35^, it was most frequent in AFR individuals. Furthermore, we discovered a more than two-fold higher frequency of homozygous carriers (*5/*5) in AFR individuals (0.43%) compared to individuals within the other groups (ranging from 0.11% AMR ancestry to 0.16% EAS ancestry). We observed that whilst the frequency of the *5 allele in the East Asian cohort (5.2%) was identical to that reported in PharmGKB^29^, it was higher than the estimate from a recent *CYP2D6* SV-focused study^3^.

#### Identical gene duplications and multiplications

*CYP2D6* duplication and multiplication SV events are key to predicting the UM category (**Supplementary Table 5**) and we identified 16,144 carriers (3.4% of the overall population). Among them, 181 were homozygous carriers, with 170 heterozygous participants carrying alternative identical gene duplication or multiplication haplotypes (e.g., *1×2/*2×2, *1×3/*2×3). Overall, we observed a higher frequency of carriers in non-EUR individuals (**Figure 3A, Supplementary Table 6**), for example, one in seven (14.5%) of AFR participants were identified as identical gene duplication or multiplication carriers. Furthermore, we found that *10+*36xN exhibited the widest range of and highest copy numbers in the EAS participants (N=1, 2, 3, 4, 5, 6, 22; **Supplementary Table 3**). Interestingly, we did not observe any multicopy carriers of the *4 allele in EAS participants, which is the most frequent star allele contributing to non-functional *CYP2D6* enzyme in EUR individuals (haplotype frequency 14%), further demonstrating the ancestry-specific distribution of *CYP2D6* star alleles.

#### Hybrid genes

Certain *CYP2D6* star alleles indicate unequal recombination between *CYP2D6* and the adjacent homologous *CYP2D7* pseudogene that create hybrid genes. The naming convention for these hybrid genes depends on the orientation of the sequence, with the gene occupying the 5’ portion mentioned first. In our cohort, we observed the currently only one *CYP2D7-2D6* hybrid allele (*13) and three *CYP2D6-2D7* hybrid alleles (*4.013, *36, *68). We found that singleton *13, *36, and *68 hybrids were rare (**Supplementary Table 3**), and we observed only one unique gene recombination arrangement for *13 (*CYP2D6*\*2+*13), *4.013 (*CYP2D6*\*4+*4.013), and *36 (*CYP2D6*\*10+*36). While, for *68, we observed three non-identical gene duplications: *1+*68, *4+*68, and *45+*68, which were most frequent in AMR (2.1×10^-3^), EUR (0.05), and AFR (6.8×10^-4^) groups, respectively. Consistent with previous findings^3^, 26.3% of *CYP2D6*\*4 European carriers had the hybrid arrangement pattern: *4+*68xN (N=1,2).

In summary, we observed that 54.6% *CYP2D6* SV carriers consisted of hybrid genes with non-identical gene duplications or multiplications. While Gustafson et. al. (2024)^36^ stated it could be challenging to identify both the gene deletion and the hybrid arrangements in the same individual for *CYP2D6* in the short read WGS, in total, we found that 2,003 participants carrying a whole gene deletion (*5) and another type of SV on different haplotypes (**Figure 3B**). Among them, 1,898 individuals had a combination of whole gene deletion (*5) and hybrid genes with non-identical gene duplication or multiplication arrangements, including the recently reported *5/*10+*36^36^. Under this genotype setting, the most frequent was *CYP2D6*\*4+*68/*5 carried by 1,533 individuals. Additionally, we found 84 individuals with the genotype combining *CYP2D6*\*5 and identical gene duplications or multiplications, including *5/*17×2, *5/*36×2, and *5/*43×2. Furthermore, we found 21 individuals had genotypes combining *CYP2D6*\*5 and singleton hybrid genes, which were *5/*13, *5/*36, and *5/*68.

### Underrepresentation of predicted rare protein-truncating variants in the current evaluation of *CYP2D6* genotypes

We used the known effects of star alleles on *CYP2D6* enzyme activity, to classify individuals into poor (PM), intermediate (IM), normal (NM), and ultra-rapid (UM)^11^ metabolisers **(Methods)**. Cohort-wide, 49.9% of individuals were predicted as NM, 39.6% as IM, 7.0% as PM and 1.6% as UM, with the remaining 1.9% as indeterminate because of the unknown effect of one or more of the identified star alleles on enzyme activity status (**Figure 4A**, **Table 1**). Across genetic ancestries, the prevalence of NM was higher in non-EUR participants, especially in AMR and SAS genetic ancestries where it represented almost two thirds of individuals.

**Figure 4.**
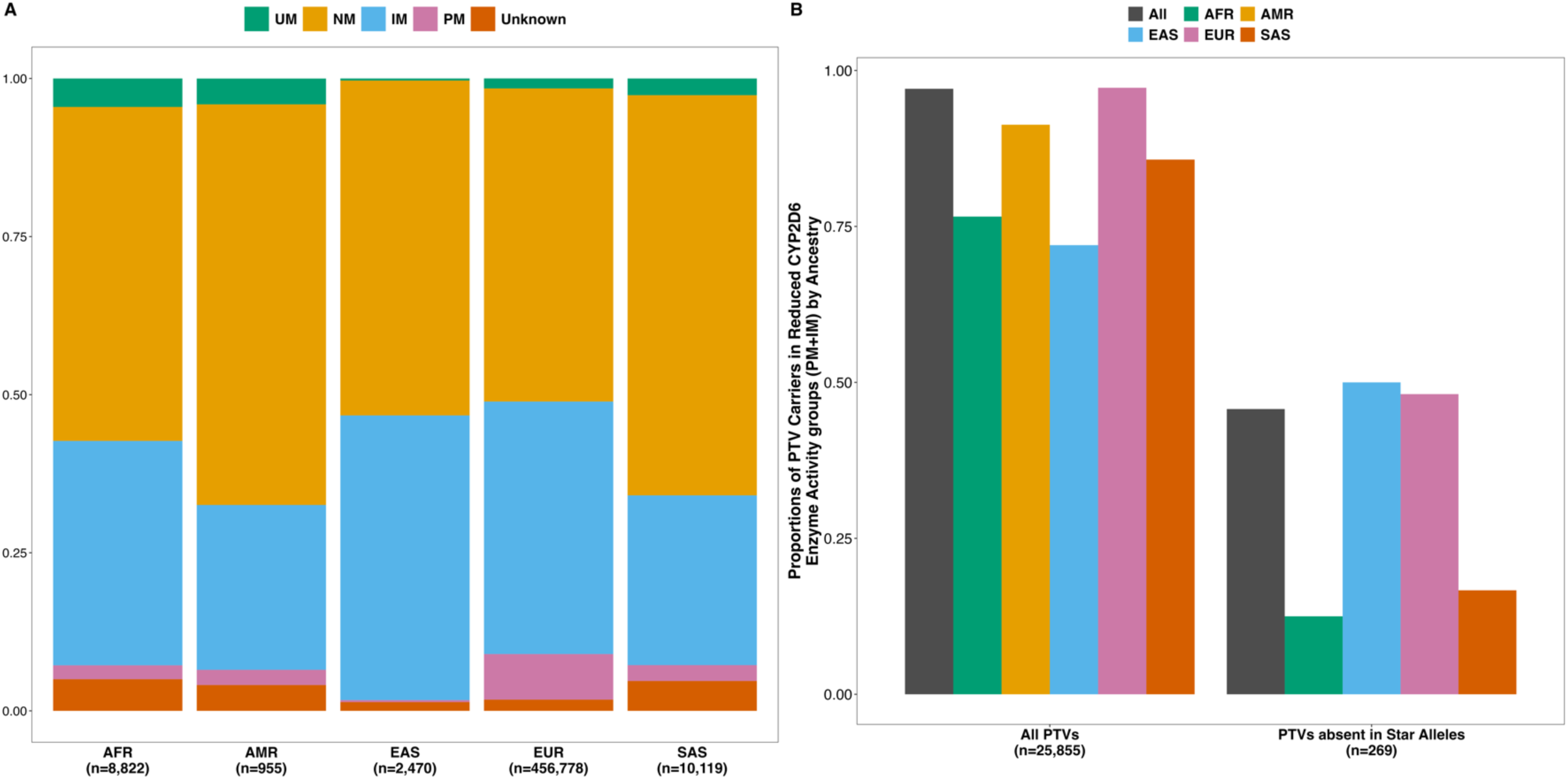
Overview of the distribution of *CYP2D6* gPhenos in the UKBB, and the comparison with CGR QV carriers in selected collapsing models. A) Distributions of *CYP2D6* gPhenos for each ancestral group in the UKBB. The bars were coloured according to the proportion of each *CYP2D6* gPheno carriers in each ancestry. Sizes of the ancestral population were included in the x-axis. B) Proportions of PTV carriers in the reduced *CYP2D6* function gPheno categories (PM or IM), coloured by genetic ancestral groups. The x-axis displayed the count of carriers of the overall *CYP2D6* PTVs (MAF≤5%) and those absent in the current *CYP2D6* star allele evaluation, respectively. The proportion on the y-axis was determined by dividing the number of PTV carriers in the decreased *CYP2D6* function gPheno categories (PM or IM) by the total number of identified carriers in the corresponding group.

**Table 1.** Overview of *CYP2D6* genetically predicted metaboliser phenotypes.

|  | **AFR** | **AMR** | **EAS** | **EUR** | **SAS** |
| --- | --- | --- | --- | --- | --- |
| **PM** | 194 (2.2%) | 23 (2.4%) | 8 (0.3%) | 32,913 (7.2%) | 254 (2.5%) |
| **IM** | 3,131 (35.5%) | 249 (26.1%) | 1,112 (45%) | 182,434 (39.9%) | 2,717 (26.9%) |
| **NM** | 4,659 (52.8%) | 605 (63.4%) | 1,308 (53%) | 226,143 (49.5%) | 6,403 (63.3%) |
| **UM** | 397 (4.5%) | 39 (4.1%) | 8 (0.3%) | 7,222 (1.6%) | 267 (2.6%) |
| **Unknown** | 441 (5%) | 39 (4.1%) | 34 (1.4%) | 8,066 (1.8%) | 478 (4.7%) |

PM – poor metaboliser; IM – intermediate metaboliser; NM – normal metaboliser; UM – ultra-rapid metaboliser; Unknown – individuals carrying star allele with currently unknown-effect. Number for each cell represents the count of individuals who were predicted to belong to the corresponding *CYP2D6* metaboliser category. The percentage in the bracket represents the proportion of the ancestral population in each *CYP2D6* metaboliser category.

We identified 78 predicted protein-truncating variants (PTVs) with a minor allele frequency (MAF) ≤ 5% in the *CYP2D6* coding regions (**Methods**, **Supplementary Table 7**), carried by a total of 25,855 individuals. Overall, we observed a significant enrichment, with 97.1% of PTV carriers showing reduced *CYP2D6* enzyme activity (i.e., categorised as PM or IM). However, this enrichment was significantly lower in the AFR (76.6%), EAS (72.0%), and SAS (85.7%) cohorts (**Figure 4B, Supplementary Table 8**). This variation may be attributed to different patterns of linkage disequilibrium between *CYP2D6* star alleles and PTVs across different genetic ancestral groups.

We found that 76 of the *CYP2D6* predicted PTVs were rare mutations with a MAF of < 0.1%. 65 of which were not included in the current evaluation of *CYP2D6* star alleles and were carried by 269 individuals. Notably, less than half (45.7%) of these people were predicted with reduced *CYP2D6* enzyme levels (i.e., PM or IM). The proportion was even lower in the AFR and SAS cohorts, at 12.5% and 16.7% respectively (**Figure 4B, Supplementary Table 8**). This indicates an underrepresentation of rare functional variants in the current *CYP2D6* star allele definition, potentially affecting the accuracy of metaboliser phenotype predictions across different genetic ancestral groups, disproportionally.

### Associations between predicted *CYP2D6* metabolisers and clinically relevant phenotypes

We sought to examine whether there were associations between *CYP2D6* gPhenos and UKBB^37^ phenotypes by performing a phenome-wide association analysis, including the plasma abundance of 2,923 proteins from the UKBB PPP data^34,38^ in a subset of 47,599 individuals. Considering the uneven distribution of predicted *CYP2D6* gPhenos, we established 12 binary pairs (**Methods, Supplementary Table 9**). These pairs involved comparisons among various combinations of *CYP2D6* metaboliser groups, including a negative control that combines PMs with UMs, contrasted with a comparison involving IMs and NMs.

Following the adjustment for multiple testing (corrected P=2×10^-7^; **Methods**), we uncovered significant associations between one or more of our *CYP2D6* gPheno contrasts and two binary clinical outcomes, six quantitative traits, and changes of three plasma protein levels (**Figure 5A&B, Supplementary Table 10-12**).

**Figure 5.**
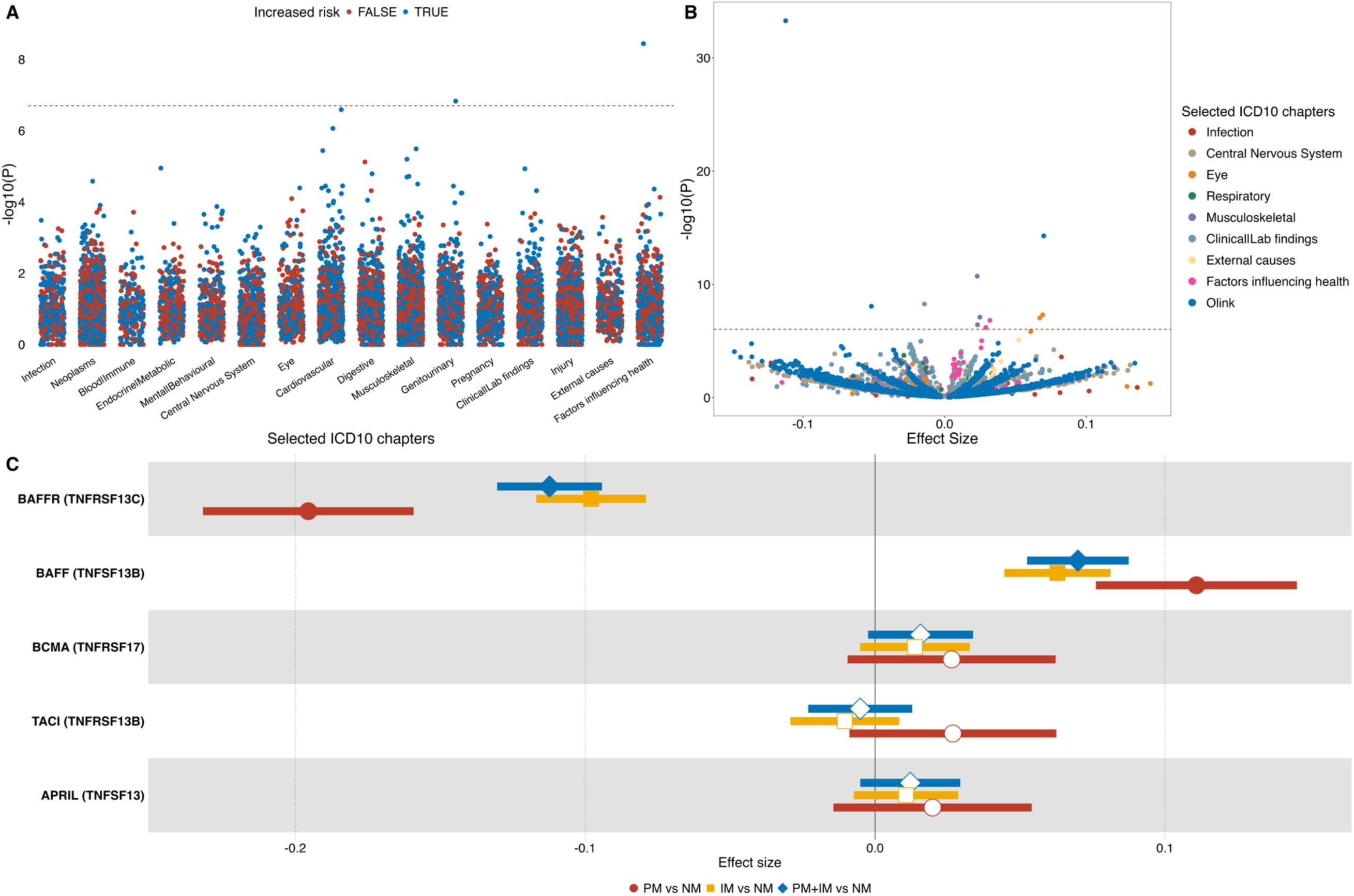
Associations of *CYP2D6* metabolisers with clinical outcomes, biomarkers, and plasma protein abundance in the UKBB. A) Manhattan plot illustrating the phenome-wide association study of binary clinical outcomes with the defined *CYP2D6* gPheno category contrasts. Each point on the plot represents a binary clinical outcome test along with the most significantly associated *CYP2D6* gPheno comparison pair. If the estimated odds ratio exceeds one (indicating individuals in the case group of *CYP2D6* gPheno(s) had higher odds of the risk), the association is marked as ’increased risk=TRUE’. The selection of the ICD10 category for presentation was based on having at least one association with a significance level below 5% (i.e., P < 0.05). The red dotted horizontal line reflects the minus log10 transformed significance threshold after the correction for multiple testing (P=2×10^-7^). B) Volcano plot illustrating the phenome-wide association studies of quantitative traits with the defined *CYP2D6* gPheno comparison pairs. Every point depicted on the plot symbolises the evaluation of quantitative traits along with the most significantly associated *CYP2D6* gPheno comparison pair. The ICD10 chapter was chosen for presentation if it had at least one association with a significance level below 5% (i.e., P < 0.05). The red dotted horizontal line reflects the minus log10 transformed significance threshold after the correction for multiple testing (P=2×10^-7^). C) Forest plot illustrating the associations between selected *CYP2D6* gPheno comparison pairs and TNF family members relevant to B lymphocyte development. The plasma protein abundances underwent a rank-based inverse normalisation transformation. The effect size estimate displayed on the x-axis indicates the change per standard deviation of the plasma protein abundance when comparing between the control and case *CYP2D6* gPheno groups. Dots were filled if the association remained significant after the correction for multiple testing (P=2×10^-7^).

Importantly, we identified a significant association between *CYP2D6* NM and UM individuals compared to PM and IM metabolisers with ‘Allergy events related to narcotic agents’ (ICD10: Z88.5, OR=1.19, 95% CI: [1.12-1.27], P=4.5×10^-9^), serving as a positive control^18^. According to the CPIC guidelines, individuals with *CYP2D6* UM status should avoid certain opioid drugs (such as codeine and tramadol) because of the risk of severe toxicity, whereas PMs should avoid these medications because of lack of efficacy^18^. Underscoring this when we limited association analysis to UM vs PM metaboliser groups, we observed a higher point estimate of the effect size (OR=1.87, 95% CI: [1.47, 2.37], P=3.5×10^-7^). Additionally, we identified an association between increased risk of developing Calculus of the kidney in NM &UM gPhenos compared to PM & IM (OR=1.19, 95% CI: [1.12, 1.27], P=8.9×10^-8^).

Quantitative trait associations included ankle spacing width (PM & IM VS NM; beta=0.02, 95% CI: [0.02, 0.03], P=1.1×10^-11^), serum creatinine (PM & IM vs NM & UM; beta=-0.01, 95% CI: [-0.02, -0.01], P=2.5×10^-9^), distance to coast (PM & IM vs NM; beta=-0.02, 95% CI: [-0.02, -0.01], P=1.5×10^-8^), spherical power (PM vs NM; beta=0.07, 95% CI: [0.04, 0.09], P=7.2×10^-8^), avMSE (PM vs NM; beta=0.06, 95% CI: [0.04, 0.09], P=1.6×10^-7^), and records in HES inpatient main dataset (PM vs NM; beta=0.03, 95% CI: [0.02, 0.04], P=3.5×10^-7^), while the molecular mechanisms underlying the identified associations remained further validation in other data resources. It is possible that some of these results may represent noise or confounding by fine-scale ancestry stratification.

Looking at the subset of 47,599 individuals with plasma proteomics data we found that compared to normal *CYP2D6* metabolisers (NM), PM and IM individuals had significantly elevated plasma levels of B-cell-activating factor receptor (BAFFR; encoded by *TNFRSF13C –* PM & IM (0) vs NM (1): beta=-0.11, 95% CI: [-0.13, -0.09], P=5.2×10^-34^), and decreased levels of its ligand BAFF (encoded by *TNFSF13B -* PM & IM (0) vs NM (1): beta=0.07, 95% CI: [0.05, 0.09], P=5.2×10^-15^). For both proteins, effect sizes estimated for PM vs NM compared to those for IM vs NM were significantly different (**Methods**; Cochran’s Q test median P_BAFFR_ = 1.9×10^-4^, P_BAFF_ = 0.043), with the former almost twice of the effect size as that of the latter (**Figure 5C, Supplementary Table 12**). In addition, the estimated effect size for PM vs IM was significantly different from that for PM vs NM in the association with plasma BAFFR levels, while at the border line of significance for that with plasma BAFF levels (**Methods**; Cochran’s Q test median P_BAFFR_=4.8×10^-3^, P_BAFF_=0.06). Given the role BAFF and BAFFR play in the long-term survival of B lymphocytes^39,40^, we assessed *CYP2D6* metaboliser status associations with autoimmune diseases and cancers. We found that PM and IM individuals had a 30% and 40% lower risk in developing chronic myeloid leukaemia and follicular non-Hodgkin’s lymphoma compared to PMs at the significance level of 0.1% and 0.9%, respectively. In addition, *CYP2D6* gPhenos were not associated with the plasma expressions of other measured TNF family members relevant to B lymphocyte development (**Figure 5C**). We found that the previously reported^38^ variant rs763882049, a pQTL for both BAFFR and BAFF, was enriched in the *CYP2D6* NMs (OR [95% CI] = 1.86 [1.84, 1.90]; P=0), with a consistent direction of effects for BAFFR and BAFF. Four *CYP2D6* star alleles were enriched for rs763882049 (**Supplementary Table 13**) including *2 where we observed a three-fold increase of prevalence for rs763882049 carriers. Analyses conditioning on the rs763882049 genotype resulted in an attenuated association of *CYP2D6* PM/IM vs NM with BAFFR (beta=-0.02, 95% CI: [-0.04, -5×10^-3^], P=0.012). Additionally, we observed that NM individuals were more likely to carry rarer protein coding pQTLs^34^ in BAFFR (*TNFRSF13C*), associated with lower abundance (**Supplementary Table 14**).

Furthermore, when contrasting PM and IM, we identified *CYP2D6* NM was negatively associated with the plasma protein abundance of Proopiomelanocortin (POMC; PM & IM vs NM; beta=-0.05, 95% CI: [-0.07, -0.03], P=8.5×10^-9^), a precursor polypeptide producing various hormones, including β-endorphin. In addition, the estimated effect size for PM vs NM was significantly different from that for IM vs NM (**Methods**; Cochran’s Q test median P-value=0.041). However, the estimated effect size for PM vs IM was not significantly different from that for PM vs NM (**Methods**; Cochran’s Q test median P-value=0.19). The average plasma POMC abundance in the *CYP2D6* UM individuals was less than that of *CYP2D6* PM population at the nominal statistical significance level (P=0.02) and was not significantly different from that of *CYP2D6* IM or NM individuals (**Supplementary Table 15**).

## Discussion

Pharmacogenomics research is pivotal to the development of personalised medicine, with *CYP2D6* emerging as a critical component, given its role in metabolising over one-fifth of the clinically prescribed drugs^1^. This study of 479,144 whole-genome sequenced UKBB individuals presents the most extensive multi-ancestry study of *CYP2D6* genetic variation to date, allowing the identification of 99,656 (20.8%) carriers of *CYP2D6* SVs.

Across the five broad genetic ancestry groups, we identified 95 distinct *CYP2D6* star alleles, with the highest number observed in EUR individuals, contrary to expectation given higher AFR genetic diversity^41^. We attributed this to the limited cohort size of non-EUR genetic ancestries in the UKBB, which may have restricted our ability to identify less common and ancestry-specific star alleles. We were unable to assign function and therefore *CYP2D6* metaboliser for 48 of the star alleles (50%), however, collectively these were found in just 1.9% of individuals. Notably, these individuals were most prevalent in AFR, AMR, and SAS individuals (4-5%) highlighting the need to identify and functionally characterise *CYP2D6* variation in these ancestries to facilitate equitable capacity to provide pharmacogenomic advice.

Comparing our findings with the published *CYP2D6* frequencies from PharmGKB, we observed high concordance in star allele haplotype frequencies for AFR, AMR, EUR, and SAS broad genetic ancestry groups (r_min_=0.97). However, concordance was lower (r=0.89) for EAS, primarily due to a discrepancy in the estimated allele frequency for *10 that might be due to the incomplete resolution of the *10+*36 haplotype in PharmGKB^29^. We also replicated an underestimation of the allele frequency of *1 for SAS genetic ancestries in PharmGKB^10^, as well as that for AFR and EUR.

*CYP2D6* SVs were observed across broad genetic ancestry groups at different frequencies, with highest rates (63.6%) observed in EAS. Overall, the most frequent (31.5%) SV was found in the *CYP2D6*\*5 (whole gene deletion haplotype), highlighting its prevalence in the population.

Whilst, ultra-rapid metaboliser (UM) phenotype invariably involved identical gene duplications or multiplications, we found that other metaboliser phenotypes could also feature such duplications, (e.g. *10+*36xN, activity value=0.25). Additionally, we found that non-identical duplications often comprised hybrid genes with ancestry-informative characteristics. Overall, these observations emphasise the need to detect *CYP2D6* SVs in clinical testing to ensure accurate therapeutic interventions and minimise potential adverse reactions especially in underrepresented genetic ancestries.

In a previous study that utilised established qualifying models developed for phenome-wide association studies (PheWAS)^32^, we identified individuals carrying computationally predicted deleterious genetic mutations affecting the functionality of their protein-coding genes. Nearly all (99%) cis-pQTL signals from the *ptv* model were linked to decreased plasma levels, indicating the reliability of variation annotations^34^. The strong enrichment (97.1%) of more common PTV variant (MAF≤5%) carriers in reduced enzyme activity groups (i.e., IM or PM) corroborated with predictions based on called *CYP2D6* star alleles. However, 65 (85.5%) rare PTVs (MAF≤0.1%) were not considered in the current *CYP2D6* star allele definition. Less than half of these rare PTV carriers (45.7%) were predicted to have reduced *CYP2D6* enzyme activity levels (i.e., PM or IM) based on the current star allele evaluation, with further reduction of the proportion in AFR (12.5%) and SAS (16.7%) cohorts, indicating the underrepresentation of the *CYP2D6* rare functional variants in the current star allele definition. Although the contribution may be marginal, incorporating further *CYP2D6* rare functional variants into star allele definitions could improve the prediction of the *CYP2D6* metaboliser activity for some individuals, especially in non-European ancestry populations. Further functional analyses are warranted to integrate newly identified rare functional variations into the expanding repertoire of *CYP2D6* star alleles.

A Phenome-wide analysis revealed the increased risk of *CYP2D6* NM and UM individuals of allergic reactions to narcotic agents, such as opioids like morphine and oxycodone, serving as a positive control. Using plasma proteome data available for a subset of approximately 50,000 individuals, we observed a significant association of *CYP2D6* PMs and IMs with the elevated level of plasma POMC, compared to NMs. POMC is a precursor polypeptide that produces β-endorphin, an endogenous opioid that binds to the µ-opioid receptor^42^. This receptor is also targeted by exogenous opioids like morphine^43^ and fentanyl^44^. *CYP2D6* is crucial for metabolising the prodrug codeine into morphine, facilitating pain relief. However, *CYP2D6* PM individuals do not effectively convert codeine, leading to reduced effect in pain relief^45^. An increase in β-endorphin among these individuals may serve as a natural compensatory mechanism. Conversely, no significant differences were found in plasma POMC expression between PM/IM/NM and UM individuals, which might be because of the considerable variation in the plasma POMC expression due to the limited size of the UM cohort. *CYP2D6* UMs, due to their heightened enzyme activity, may produce excessive morphine, potentially leading to toxic effects after codeine ingestion.

Furthermore, studying the UKBB PPP data enabled us to identify significant *cis*- and *trans*-associations between *CYP2D6* metaboliser phenotypes and plasma levels of BAFF-R and BAFF, respectively. The *cis-*association with BAFFR might be attributed to the linkage between the previous identified^38^ BAFFR *cis-*pQTL (rs763882049) with certain *CYP2D6* star alleles. BAFF is a well-known target for belimumab, which has been approved for the clinical treatment of systemic lupus erythematosus^46^. Although no direct associations were found between *CYP2D6* metaboliser phenotypes and common autoimmune diseases in the UKBB dataset, our findings suggest a potential gene-drug relationship between *CYP2D6* and BAFF-targeted autoimmune clinical treatments, warranting further dosage optimisation experiments tailored to different *CYP2D6* metaboliser phenotypes, and highlighting the potential need of *CYP2D6* metaboliser level assessment prior to the BAFF-targeted clinical practice.

We acknowledge several limitations to our study. Firstly, the UKBB dataset is predominantly enriched in EUR participants (95.3% in our study), limiting our ability to fully survey ancestry informative *CYP2D6* genetic features for non-EUR cohorts. However, our study highlights the opportunities to deliver more equitable access to medicines through the growing availability of large scale whole genome sequencing data of non-European broad genetic ancestry^47^. We chose not to study the relationship between *CYP2D6* genetic variation and clinical prescription data in the UKBB as these data were only available for a subset of individuals (45.2%), there is an average five-year interval between phenotype measurements, and there is an absence of data concerning detailed consequences for changes in drug types and dosages, which complicates modelling the efficacy of medication and dosage changes, as well as the associated occurrences of side effect over time. Finally, functional analyses will be required to validate the impact on enzyme function for the novel *CYP2D6* variants that we describe and how these might affect clinically relevant phenotypes.

In summary, our study, is to our knowledge, the largest to date to employ WGS data to analyse *CYP2D6* pharmacogenomics across multiple ancestries. We highlight the lack of genetic ancestral diversity in the current evaluation of *CYP2D6* star alleles and emphasise the importance of including structural variations, available through WGS, to accurately predict *CYP2D6* metaboliser phenotypes. Our results reveal clinically significant associations that enhance the understanding of molecular mechanisms underpinning previously identified drug responses and suggest potential dosage optimisation for BAFF-targeted treatments.

## Data and code availability

The UK Biobank data were available via the registration for access procedure described at https://www.ukbiobank.ac.uk/enable-your-research. Association tests described in this study were performed using a custom framework, PEACOK (PEACOK 1.0.7), that is available via GitHub (https://github.com/astrazeneca-cgr-publications/PEACOK/).

## Acknowledgments

We thank the participants and investigators in the UKB study who made this work possible (Resource Application Number 26041). We are grateful to the research and development leadership teams at the 13 participating UKB-PPP member companies (Alnylam Pharmaceuticals, Amgen, AstraZeneca, Biogen, Bristol-Myers Squibb, Calico, Genentech, Glaxo Smith Klein, Janssen Pharmaceuticals, Novo Nordisk, Pfizer, Regeneron and Takeda) for funding the study.

## Author contributions

X.J. and K.R.S conceptualised and designed this study. X.J and F.H. performed analyses and statistical interpretation. X.Z.Z. and S.S.A. conducted the structural variation analyses using an independent calling pipeline. S.V.V.D did the bioinformatics processing. A.A., G.A., A.O., J.H., and M.F. contributed to biological interpretation. X.J., O.B., and K.R.S. drafted the main text and supplementary materials. Q.W., S.P., and W.R. supervised the study. All authors read, commented on, and agreed upon the submitted manuscript.

