## Supplementary Figures for "Genetic ancestral patterns in *CYP2D6* alleles: structural variants, rare variants, and clinical associations in 479,144 UK Biobank genomes"

**Supplementary Figure 1. Comparisons in frequencies of *CYP2D6* star allele haplotype between the UKBB ancestries and the matching biogeographic groups in PharmGKB**

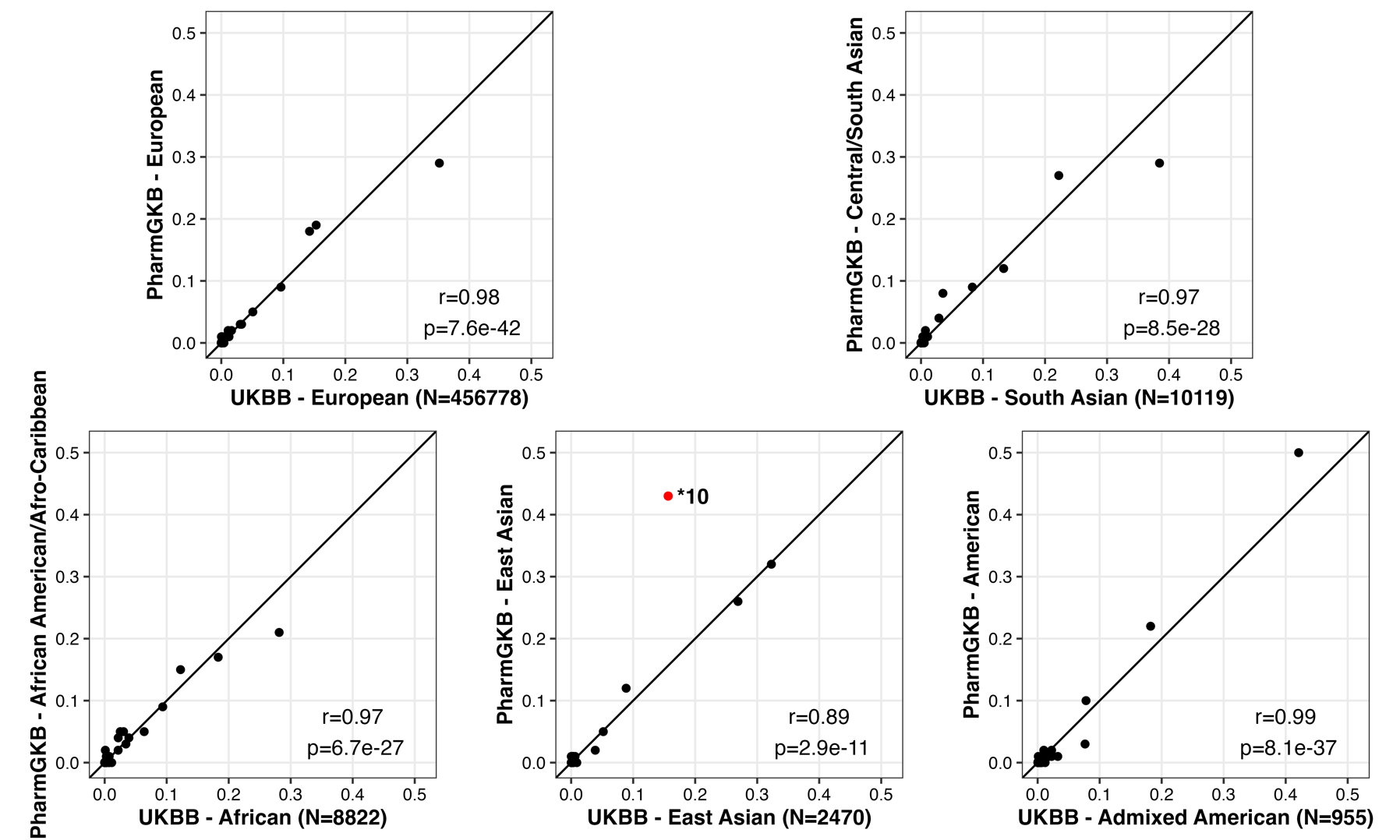

This figure depicts the comparisons of *CYP2D6* star allele haplotype frequencies between the corresponding biogeographic groups from UK Biobank (UKBB) and PharmGKB, presented individually. The frequencies were largely consistent across the European, South Asian, African, and admixed American cohorts in both datasets. However, the correlation in East Asian ancestry exhibited a relatively lower agreement, primarily influenced by the *10 allele (highlighted in red).

**Supplementary Figure 2.** **Comparisons in frequencies of *CYP2D6* star allele haplotype between the African cohort in the UKBB and that of the ‘Sub-Saharan African’ group in PharmGKB**

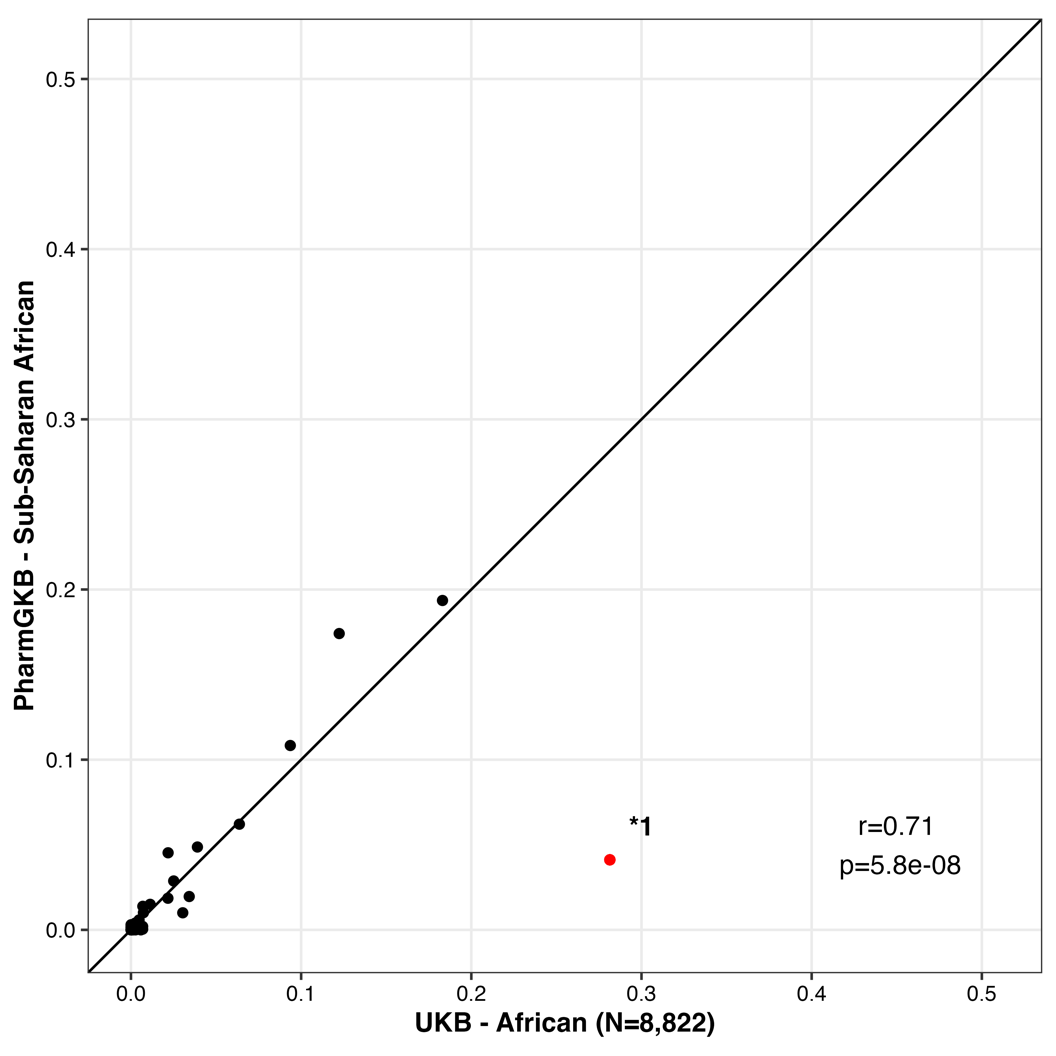

The figure demonstrates the comparisons of *CYP2D6* star allele haplotype frequencies between the African cohorts from the UK Biobank (UKBB) and the "Sub-Saharan African" cohort in PharmGKB resources. The correlation between the two datasets was impacted significantly by the notable difference in the frequencies of the *1 allele.
